# Alzheimer’s and Parkinson’s diseases predict different COVID-19 outcomes, a UK Biobank study

**DOI:** 10.1101/2020.11.05.20226605

**Authors:** Yizhou Yu, Marco Travaglio, Rebeka Popovic, Nuno Santos Leal, L. Miguel Martins

## Abstract

In December 2019, a coronavirus, severe acute respiratory syndrome coronavirus 2 (SARS-CoV-2) began infecting humans causing a novel disease, coronavirus disease 19 (COVID-19). This was first described in the Wuhan province of the People’s Republic of China. SARS-CoV-2 spread throughout the world causing a global pandemic. To date, thousands of cases of COVID-19 were reported in the United Kingdom, and over 45,000 patients have died. Some progress has been achieved in managing this disease, but the biological determinants of health, besides age, that affect COVID-19 infectivity and mortality are under scrutiny. Recent studies show that several medical conditions, including diabetes and hypertension, increase the risk of COVID-19 infection and death. The increased vulnerability of the elderly and those with comorbidities, together with the prevalence of neurodegenerative diseases with advanced age, led us to investigate the links between neurodegeneration and COVID-19. We analysed the primary health records of 13,338 UK individuals tested for COVID-19 between March and July 2020. We show that a pre-existing diagnosis of Alzheimer’s disease predicts the highest risk of COVID-19 infection and mortality among the elderly. In contrast, Parkinson’s disease patients were found to be at increased risk of infection but not mortality from COVID-19. We conclude that there are disease-specific differences in COVID-19 susceptibility among patients affected by neurodegenerative disorders.

## INTRODUCTION

The rapid emergence of coronavirus disease 19 (COVID-19) has caused over one million deaths worldwide ^1^. The clinical features of patients affected by COVID-19 have been extensively explored, but the predisposing factors contributing to increased transmission and clinical severity remains unclear. Social and ecological health determinants such as air pollution ^2-5^, have been suggested to increase the risk of infection and exacerbate COVID-19 related illness. In addition, several comorbidities have been shown to increase COVID-19 mortality rates, including cardiovascular and respiratory pathologies ^6,7^. The predominance of these comorbidities in advanced age combined with the increased incidence of mortality in elderly patients indicates that age is a major risk factor for COVID-19 mortality ^8^. These observations raise concerns that elderly patients affected by chronic diseases, including neurological manifestations, may be susceptible to increased risk of critical illness from COVID-19. In addition to COVID-19, age is one of the major risk factors for neurodegenerative disorders such as Alzheimer’s disease (AD) and Parkinson’s disease (PD)^9^. Moreover, cardiovascular and respiratory pathologies are not only risk factors for COVID-19 mortality, but also for dementia ^10-12^. In fact, new evidences show that pre-existing diagnosis of dementia represents an important risk factor for COVID-19 ^13^. In one of the largest cohort studies of COVID-19 in Europe, data gathered from 166 hospitals in England, Scotland, and Wales showed that dementia was amongst the most common comorbidities in the 20,133 patients hospitalised for COVID-19, after adjusting for age and other confounders ^14^. Another study showed that patients with dementia and other neurological diseases are at higher risk of COVID-19 in-hospital death ^15^. In addition, an analysis of 153 non-demented PD patients, without a history of lung disease, found that 40% of these patients had respiratory difficulties ^16^, showing that some sub-groups of PD patients may be at elevated risk of severe complications following severe acute respiratory syndrome coronavirus 2 (SARS-CoV-2) infection. Similarly, the high incidence of cognitive impairment in patients with AD, particularly in the late stages of the disease, might be an important risk factor for COVID-19 infection and mortality ^17^. These studies provide compelling evidence that individuals with dementia or neurodegenerative diseases are at higher risk of COVID-19 infection and mortality, but it remains unclear if specific subtypes of neurological disorders may differentially affect COVID-19 outcome. Furthermore, it remains unclear which mechanisms underlie this putative association and whether specific types of neurodegenerative diseases are associated with increased COVID-19 infection and mortality.

A recent review indicates that disease-specific neuropathology may underlie differential vulnerability to COVID-19 infection and mortality among individuals affected by AD or PD ^17^. In England and Wales, 25.6% of all COVID-19 deaths registered between March and June 2020 are found in people with dementia, making these the most common pre-existing conditions associated with COVID-19 mortality ^18^. However, it remains unclear if patients with PD are at greater risk of COVID-19 adverse outcome compared to the general population ^19^. Most studies to date only included data from hospitalised or clinically suspected COVID-19 patients. Because a large proportion of COVID-19 infections are asymptomatic ^20^, these inclusion criteria likely led to some degree of misclassification, as hospital admission rates for COVID-19 depend on the prevalence of community testing and admission criteria, which vary between countries.

Here we explored whether individuals with dementia, AD or PD are at increased risk of COVID-19 infection and mortality using the UK Biobank. This dataset contains primary health records of more than half a million individuals, including participants that were tested for COVID-19 since the beginning of this pandemic. We found that a pre-existing diagnosis of dementia or AD predicts the largest risk of COVID-19 infection and mortality. PD was associated with increased risk of infection but not mortality from COVID-19. Findings from previous studies suggested an important role for dementia in increasing susceptibility to COVID-19 ^14^. Here we provide the first systematic analysis of the relationship between COVID-19 and neurodegenerative diseases at the individual level. Our results have important implications for disease management and further highlight an important role of disease-specific neuropathology and management in the potential susceptibility to COVID-19.

## METHODS

### UK Biobank data sources

The UK Biobank comprises health data from over 500,000 community volunteers aged 40 to 70 years at baseline (2006 to 2010), living close to 22 assessment centers in England, Scotland and Wales ^21^. Baseline assessments included demographics, lifestyle and disease history, with linkages to electronic medical records. UK Biobank ethical approval was granted from the North West Multi-Centre Research Ethics Committee. The current analysis was approved under the UK Biobank application #60124. Details regarding the geographical regions, recruitment processes, and other characteristics have been previously described ^21^. A detailed list of the variables analysed in the present study is presented in Supplementary Table 1. We defined hypertension using the criteria of a diastolic blood pressure ≥ 90 mmHg OR systolic blood pressure ≥140 mmHg. Individual-level data was collected from the UK Biobank on August 17, 2020.

### Study design and exclusion criteria

We conducted a cohort study using national primary care electronic health record data linked to in-hospital COVID-19 death data (see UK biobank data sources). Of the 13,338 participants with available COVID-19 data, 1,626 tested positive for COVID-19 between March 16 and July 26 (2020) and 11,712 were negative. The majority of samples tested for COVID-19 disease were derived from combined nose/throat swabs and analysed by real-time polymerase chain reaction (PCR). More information on the testing procedure can be found on the UK Biobank website ^22^. During the same period, COVID-19 testing in England was restricted to hospitalized patients with clinical signs of the disease and health care workers. In contrast, all UK Biobank participants included in this study were subjected to COVID-19 testing since the beginning of the pandemic. For our models, we defined a positive outcome as either a positive COVID-19 diagnosis or an in-hospital death in COVID-19 positive cases. Risk factors and covariates used for the present analysis were selected on the basis of clinical interest and prior findings. These are shown in Supplementary Table 1 and included dementia, AD, PD, frontotemporal, vascular dementia, cancer, diabetes, high blood pressure, blood group, age, sex, obesity, respiratory difficulties [Chronic obstructive pulmonary disease (COPD) and wheezing], forced expiratory volume (FEV), gray and white matter volume, brain volume, white matter hyperintensity and C-reactive protein levels (CRP). Obesity was defined based on waist-to-hip ratio measurements. Waist-to-hip ratio is determined by dividing waist circumference by hip circumference, meaning that overweight individuals have higher ratios ^23^. We grouped smoking status into current, former and never smokers. CRP total levels were normalised according to protein total protein levels, and log-transformed to fit a normal distribution. Other covariates considered as potential upstream risk factors were population density, social deprivation, average household income, education level, housing type, ethnicity, environmental risk at workplace (chemical, diesel, dusty, smoke), travel to work and number of people per household. Deprivation was measured using the Townsend Social Deprivation Index (TSDI) ^24^, with higher values indicating higher deprivation, derived from the patient’s postcode for a higher degree of precision. Ethnicity was grouped into white or minority ethnicities. The full list of minority groups used for our analysis can be found in Supplementary Table 2. Information on all covariates were obtained from primary care records provided by the UK Biobank. The geographical distribution of each subject included in the analysis is shown in Figure 1.

**Figure 1.**
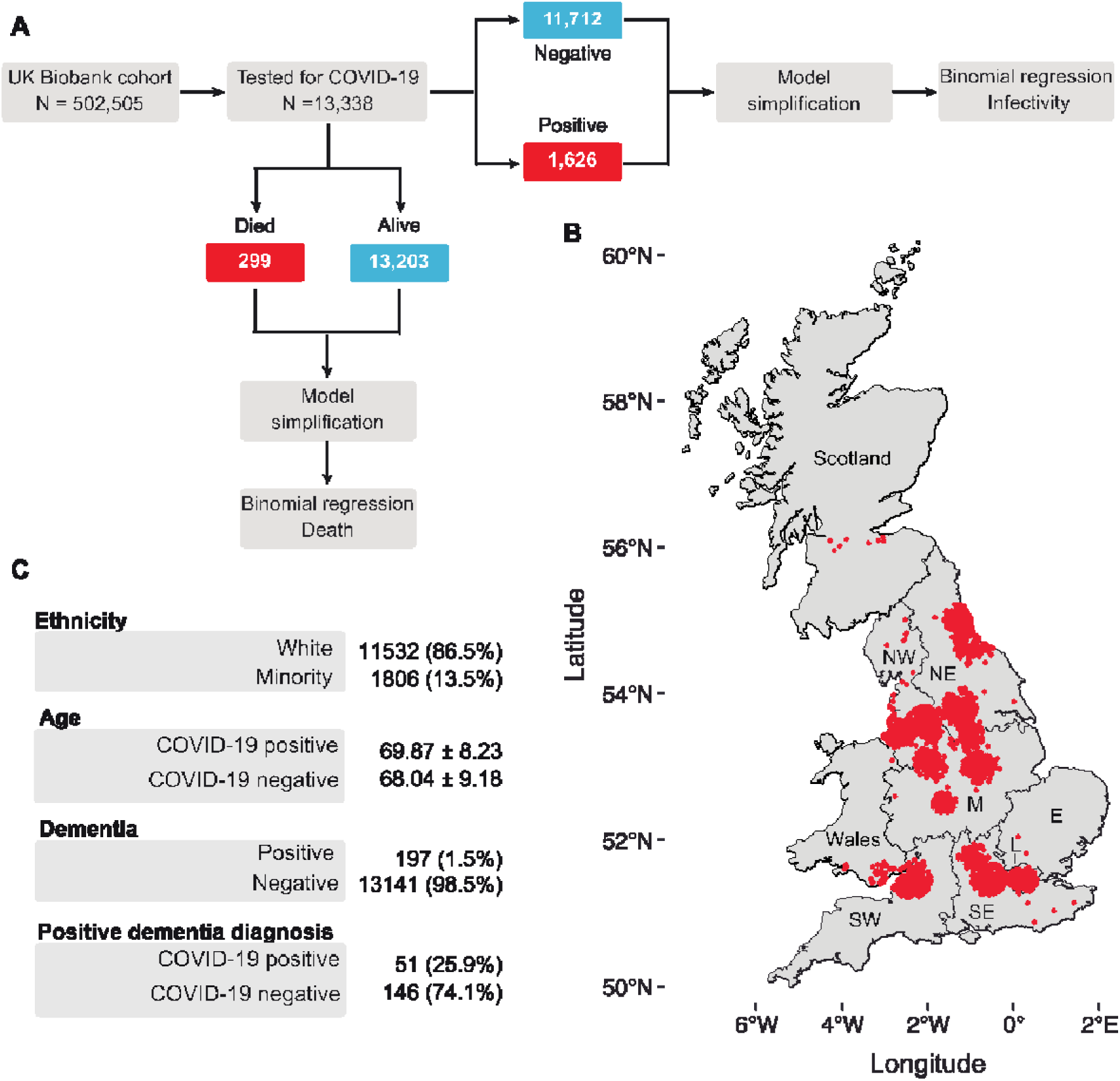
Summaries of the workflow and statistics of the analysed cohort. **A**, Workflow of the analysis. Of 505,505 UK Biobank participants, 13,338 participants were tested for COVID-19 as of 26 July 2020. We modelled the covariates that affect COVID-19 infectivity or COVID-19-related death using only participants that were tested for COVID-19. We define a COVID-19-related death as a participant who both tested positive for the COVID-19 virus and died. **B**, Distribution of the UK Biobank participants in the United Kingdom. **C**, Descriptive statistics of the cohort analysed. The number of participants in each category precede their percentage with respect to the total cohort. For the “Age” category, the mean and standard deviation are shown. A full table of summary characteristics is available in Supplementary Tables 1 and 2. SW, South West; SE, South East; E, East; M, Midlands; NE, North East; NW, North West.

### Statistical analysis

For the analysis of COVID-19 infection and mortality, we fitted binomial regression models where the response variables were, respectively, COVID-19 positivity or COVID-19-related death. We defined COVID-19-related death as an individual who tested positive for COVID-19 and died. For the multivariable model of COVID-19 infection, we omitted FEV, COPD, environmental risk at the workplace, and travel to work due to a large number of individuals having missing data. Methods for assessing the fit of the model included residual analyses, diagnostic tests, and information criterion fit statistics. The goodness of fit of each regression model was determined using the log-likelihood and Akaike Information Criterion (AIC) statistics. When analysing COVID-19 outcome, we applied an iterative variable selection procedure combining unsupervised stepwise forward and stepwise backward regression analyses to select the most suitable predictor or combination of predictors in our models, based on AIC values. For all models, we calculated the odds or risk ratios and their 97.5% confidence intervals to quantify the effects of the independent variables on the response variables. The models were built using the MASS package ^25^ in R. The comparison tables were generated using the Stargazer package ^26^. The analysis source code, detailed quality checks as well as all Supplementary material is available in GitHub (https://m1gus.github.io/AD_PD_COVID19/). Statistical significance was defined as *p* ≤ 0.05.

## RESULTS

### AD and PD diagnoses are associated with an increase in SARS-CoV2 infections in the UK Biobank cohort

To explore the links between neurodegenerative diseases and COVID-19, we first estimated the risk between COVID-19 and chronic diseases. Chronic diseases often coexist in older adults ^11,12,27^. Therefore, we assessed their risk after adjusting for other existing comorbidities (see Figure 1A for the workflow) for an aged and predominantly white cohort in Great Britain (Figure 1B and 1C). Using an iterative variable selection procedure, combining unsupervised stepwise forward and backward regression analyses, we found that a pre-existing diagnosis of dementia is associated with the largest increase in the likelihood of testing positive for COVID-19 (OR 3.25; 97.5% CI 2.73-3.87), Supplementary Table 3. This is followed by increasing waist-to-hip ratio (OR 3.07; 97.5% CI 2.26-4.17), poor education (OR 1.67; 97.5% CI 1.38-2.02), number of people per household (OR 1.05; 97.5% CI 1.03-1.07) and social deprivation (TSDI) (OR 1.03; 97.5% CI 1.02-1.04) (Figure 2A). Our findings confirm previous studies showing that dementia predicts one of the highest risks of COVID-19 infection in elderly individuals ^28,29^. Furthermore, consistent with previous observations ^30^, our primary analysis identified white ethnicity (OR 0.72; 97.5% CI 0.66-0.77), cancer (OR 0.81; 97.5% CI 0.74-0.89) and decreasing age (OR 0.97; 97.5% CI 0.97-0.97) as significant predictors of SARS-CoV-2 infectivity, after adjusting for multiple confounding factors (Figure 2A). As recent studies highlighted links between the global burden of dementia and COVID-19 death^28^, we next assessed whether diagnosis of dementia increases the risk of COVID-19 mortality in the UK Biobank. Similar to the COVID-19 infection models, we found that a diagnosis of dementia was associated with the largest risk of mortality from COVID-19 (OR 4.32; 97.5% CI 3.33-5.60), followed by male gender (OR 1.44; 97.5% CI 1.20-1.73), increasing age (OR 1.09; 97.5% CI 1.08-1.07), and social deprivation (OR 1.07; 97.5% CI 1.05-1.09) (Figure 2B and Supplementary Table 4). Cancer was negatively associated with an increased risk of mortality in our cohort (OR 0.56; 97.5% CI 0.44-0.72).

**Figure 2.**
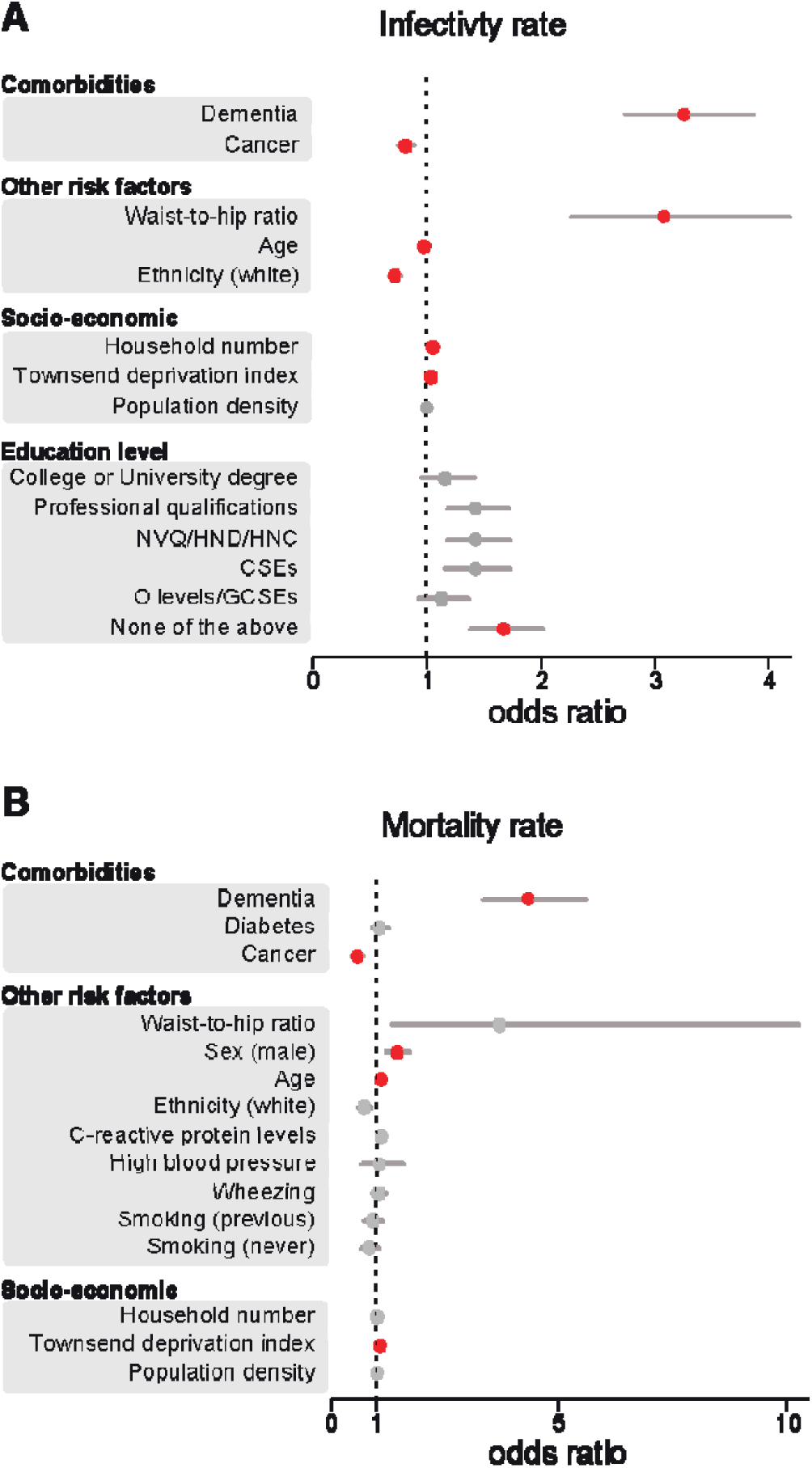
Dementia is the largest risk factor COVID-19 infectivity and death. Odds ratio and their respective 97.5% CIs for the relationship between individual-level characteristics and COVID-19 cases (**A**) or deaths (**B**). For simplicity, characteristics were sub-grouped into three categories, namely ‘comorbidities’, ‘other risk factors’ and ‘socio-economic’. Red indicates significant associations (*p* ≤ 0.05), while grey indicates a lack of significance (*p*> 0.05). The odds ratios for education levels are relative to A-levels. NVQ/HND/HNC, participants who received vocational qualifications such as National Vocational Qualifications (NVQ), Higher National Certificate (HNC) or Higher National Diploma (HND); CSEs, participants with a Certificate of Secondary Education (CSEs); O-levels/GCSEs, participants with either a General Certificate of Secondary Education (GCSE) or a General Certificate of Education (GCE) Ordinary Level (O-levels), a secondary school leaving qualification. This figure is related to Supplementary Tables 3 and 4.

Given the prominent role of dementia in COVID-19 diagnosis, we next analysed how different types of dementia are associated with COVID-19. The overall cumulative incidence of PD and AD diagnosis among COVID-19 test positive patients in our cohort was 1.7% and 1.6%, respectively (Supplementary Table 5). Our results show that a diagnosis of AD was strongly associated with COVID-19 infectivity, with AD patients showing the greatest susceptibility to COVID-19 infectivity compared to individuals without AD (OR 4.15; 97.5% CI 3.22-5.34). We also found that an increase in waist-to-hip ratio (OR 3.08; 97.5% CI 2.27-4.19) or pre-existing vascular dementia (OR 2.51; 97.5% CI 1.69-3.71) are also positively associated with COVID-19 infection (Figure 3A). PD diagnosis also emerged as a strong positive predictor of COVID-19 infection (OR 1.71; 97.5% CI 1.37-2.13), although the effect was smaller than the one for AD diagnosis. Low education level (OR 1.65; 97.5% CI 1.36-2.0), higher number of people per household (OR 1.05; 97.5% CI 1.03-1.07) or increased Townsend deprivation index (OR 1.03; 97.5% CI 1.02-1.04) emerged as significant predictors of a positive COVID-19 diagnosis (Figure 3). Our analysis shows that patients of white ethnicity (OR 0.72; 97.5% CI 0.66-0.77) or with pre-existing diagnosis of cancer (OR 0.81; 97.5% CI 0.72-0.88) are at lower risk of infection in our cohort, while increasing age does not predict increased risk of infection (OR 0.97; 97.5% CI 0.97-0.97; Figure 3A).

**Figure 3.**
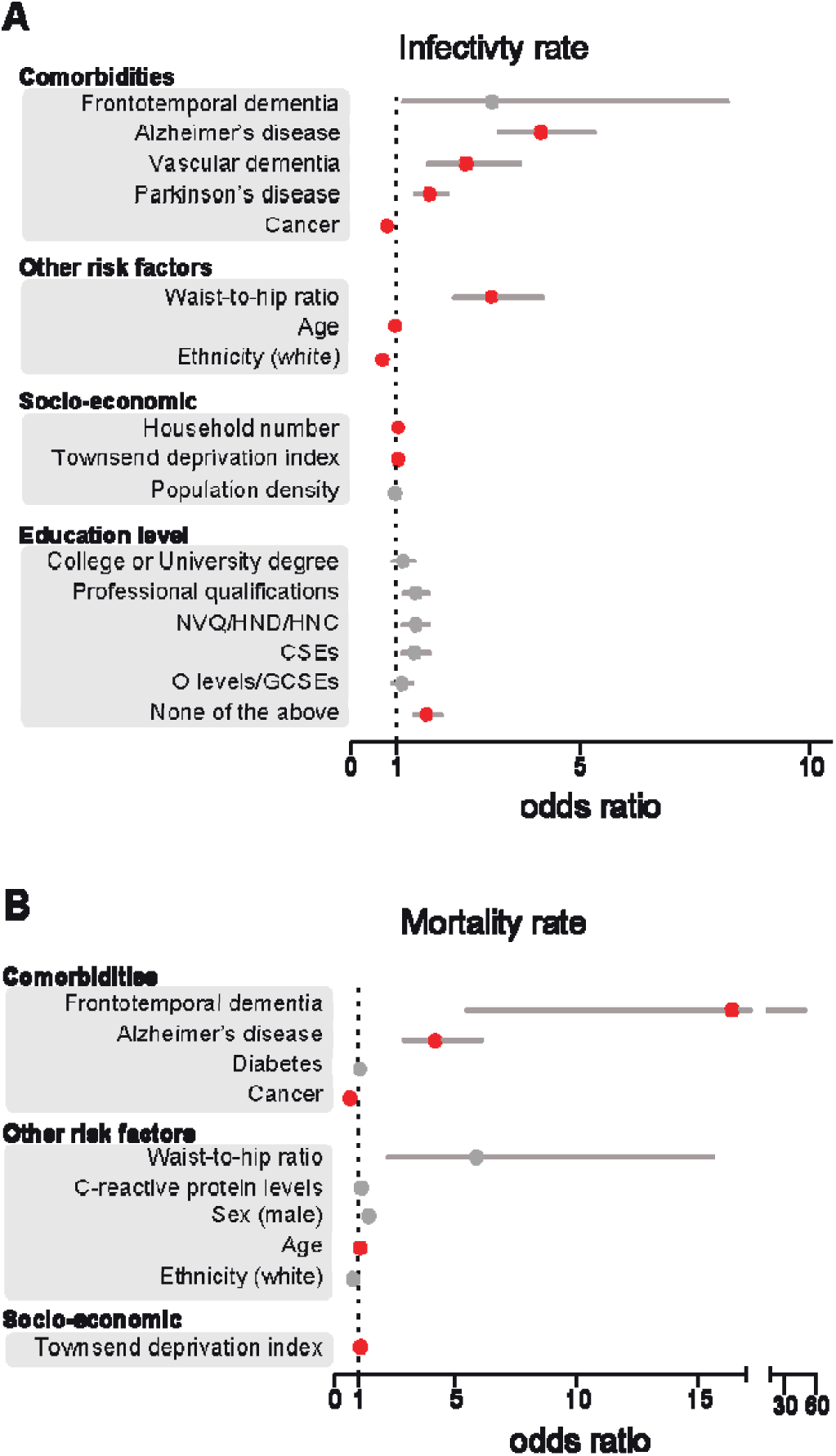
AD diagnosis predicts increased risk of COVID-19 mortality and infection. Odds ratio and their respective 97.5% CIs for the relationship between individual-level characteristics and COVID-19 cases (**A**) or deaths (**B**). For simplicity, characteristics were sub-grouped into three categories, namely ‘comorbidities’, ‘other risk factors’ and ‘socio-economic’. Red indicates significant associations (*p*≤0.05), while grey indicates a lack of significance (*p*> 0.05). The odds ratios for education levels are relative to A-levels. NVQ/HND/HNC, participants who received vocational qualifications such as National Vocational Qualifications (NVQ), Higher National Certificate (HNC) or Higher National Diploma (HND); CSEs, participants with a Certificate of Secondary Education (CSEs); O-levels/GCSEs, participants with either a General Certificate of Secondary Education (GCSE) or a General Certificate of Education (GCE) Ordinary Level (O-levels), a secondary school leaving qualification. The numeric values of this figure are in Supplementary Tables 5 and 6.

## AD patients are at higher risk of COVID-19 death

Although our results show that patients with dementia are at increased risk of contracting COVID-19, it remains unclear if the presence of neurological comorbidities may exacerbate the risk of mortality in COVID-19 patients. To address this issue, we examined the characteristics and outcomes of all COVID-19 patients in the cohort using a binary multivariable regression model (Figure 3B and Supplementary Table 6). Given the large number of variables in our models, we simplified our analysis using an iterative stepwise regression approach to select the most suitable predictors of COVID-19 death based on AIC. In terms of neurological diseases, diagnosis of frontotemporal dementia (OR 16.36; 97.5% CI 5.44-49.15) and AD (OR 4.17; 97.5% CI 2.87-6.05) were associated with the largest risk of COVID-19 death but not PD or vascular dementia. In our model, pre-existing diagnosis of cancer was negatively associated with COVID-19 death (OR 0.63; 97.5% CI 0.51-0.79) and no significant association was found between diabetes, C-reactive protein levels and ethnicity (Figure 3B). We also found that living in socially deprived areas increases the risk of COVID-19 adverse outcome (OR 1.07; 97.5% CI 1.05-1.09), while an increased waist-to-hip ratio (OR 5.83; 97.5% CI 2.18-15.58) or male gender (OR 1.38; 97.5% CI 1.16-1.65) were positively associated with COVID-19 death but this relationship did not reach statistical significance.

We next focused on the role of AD on COVID-19-related deaths. We first built a model that only contains participants with a positive AD diagnosis (Supplementary Table 7). Using this model, we found that none of the previously mentioned comorbidities were significant. This observation indicates that participants with AD are at higher risk of dying from COVID-19, independently from age, sex and other comorbidities.

## DISCUSSION

Despite considerable uncertainty in estimates of COVID-19 outcomes, age and comorbid medical conditions are consistently associated with adverse health outcomes in hospitalised COVID-19 patients ^15^. The incidence of neurological conditions, including dementia and neurodegenerative diseases, increases with age, and it has recently been proposed that individuals with pre-existing diagnosis of dementia may be at increased risk of developing COVID-19 ^13^. In previous viral outbreaks of respiratory pathogens, including severe acute respiratory syndrome, Middle East respiratory syndrome, and H1N1 influenza, several reports also highlighted the presence of neurological comorbidities in affected patients ^31,32^. In the present study, we found that the largest risk factors associated with COVID-19 infection are a pre-existing diagnosis of dementia associated with AD, vascular dementia or PD. However, while diagnosis of AD also predicts increased risk of COVID-19 mortality, our findings suggest that COVID-19 mortality among PD and vascular dementia patients does not differ from the general population.

A close relationship between COVID-19 and neurological disorders is well established in the literature. An early report ^13^ this year showed that dementia diagnosis is associated with the largest increase in the risk of COVID-19 infection in a smaller cohort of patients in England aged 65 and older. Dementia has also been found to increase the risk of in-hospital mortality in a large cross-sectional analysis of 20,133 patients already hospitalised for COVID-19 in the UK ^14^, a finding later replicated by two later studies ^15,33^. However, these studies mostly focussed on hospitalised individuals and did not include data on patients managed in the community settings, such as domestic residences. In addition, in line with government guidelines, testing was limited to individuals with COVID-19 symptoms meaning that their figures do not include the growing number of people who are asymptomatic or are self-isolating at home due to mild symptoms ^14^. Using granular data from the UK Biobank, we have developed a more robust analysis pipeline, since all participants in our dataset received COVID-19 testing. Because a large proportion of COVID-19 infections are asymptomatic, this screening protocol is more sensitive for the analysis of COVID-19 infection and mortality rates among people diagnosed with neurological diseases.

Our results indicate that AD patients are at increased risk of COVID-19 infection. These findings expand a recent analysis with a smaller sample size of COVID-19 positive individuals from the UK Biobank. In this study, Zhou and colleagues showed that AD is the most significant risk factor for COVID-19 infection but its association with increased COVID-19 mortality was not investigated ^34^. Here, we build on these earlier observations to show that AD is a major risk factor associated with COVID-19 mortality, after accounting for a large number of comorbidities. Several features of AD may increase the risk of COVID-19 adverse outcome. First, the neuropathology of AD could facilitate COVID-19 complications. Accumulating evidence suggests that amyloid fibrils induce microglial activation and increased activation of the type-1 interferon (IFN) pathway, a crucial component of COVID-19 infection ^35^. Current theories propose that the IFN response in AD may synergise with COVID-19 upon SARS-CoV-2 infection creating the ‘perfect storm’ of excessive immune responses and thus exacerbating pathology ^36^. Supporting the hypothesis of a neurobiological link between AD and COVID-19 mortality, a recent study further demonstrated that protein expression levels of angiotensin□converting enzyme 2 (ACE2), the entry receptor for SARS-CoV-2, are upregulated in the brain of AD patients ^37^. This suggests that higher ACE2 expression may underscore higher viral load in the brain of AD patents, corroborating a potential link between AD neuropathology and COVID-19 mortality ^38^. Finally, the social behaviour of patients with dementia and AD must be considered. Cognitive decline may compromise the ability of individuals with AD to follow the recommendations of public health authorities, increasing the likelihood of contagion and the need for carers ^39^. Behavioral and psychological symptoms (BPSD) of dementia and AD, such as motor agitation, intrusiveness, or wandering, may further undermine efforts to maintain isolation.

We also found that PD is associated with a heightened risk of COVID-19 infectivity but not mortality. This is consistent with a retrospective study conducted in Japan where patients with parkinsonism hospitalised with pneumonia were found to display a lower rate of in-hospital mortality than age- and sex-matched controls ^40^. Two recent studies from Italy have reported increased COVID-19 mortality rates among PD patients. One group gathered clinical information on 120 community-dwelling PD patients and reported a mortality rate of 20%, a value significantly higher than the general population ^41^. The second study found that PD patients of older age (>78 years) display increased susceptibility to COVID-19 death compared to younger patients ^42^. However, both studies used clinically suspected (non-laboratory confirmed) COVID-19 cases, which complicates their interpretation. As noted by the authors, the increased susceptibility to COVID-19 may have resulted in some patients being incorrectly identified as COVID-19 positive, thus leading to misclassification of COVID-19-related deaths. Moreover, the accuracy of prevalence data might further be hampered by the existence of asymptomatic cases and the lack of population screening campaigns in Italy. Our results support a recent case-controlled study, which shows that PD is not associated with any apparent risk of morbidity and mortality compared to the general population ^43^.

Although the biological basis for the higher mortality rate in AD compared to PD patients remains to be elucidated, it has recently been suggested that PD neuropathology itself might exercise a neuroprotective effect against COVID-19. For instance, SARS-CoV-2 binds to the ACE2 receptor, which is highly expressed in the dopaminergic neurons of the striatum ^44^. However, PD-related neuropathology induces significant degeneration of these neurons, pointing to reduced neuroinvasion in these patients ^17^. Secondly, increased neuronal expression of α-synuclein following acute West Nile virus infection suggests that this protein could function as a native antiviral factor within neurons ^45^. Finally, a number of PD drugs have been hypothesised to play a therapeutic role in COVID-19. Among these, accumulating evidence shows that amantadine may inhibit viral replication and protect against severe outcomes in PD patients ^46^. The proposed mechanism of action involves disruption of the lysosomal machinery needed for viral replication ^47^ and there is preliminary evidence of a protective effect against COVID-19 in a small cohort of PD patients, all taking L-DOPA and having tested positive for SARS CoV-2 ^48^. In the present study, none of the PD patients receiving amantadine treatment developed severe complications from COVID-19 and only one patient tested positive for SARS CoV-2 (Supplementary Table 9). Although limited by a small sample size, our preliminary analysis is in line with the hypothesis that amantadine may exert a protective effect against both COVID-19 infection and mortality. Further clinical studies should be conducted to corroborate the therapeutic utility of amantadine for the treatment of COVID-19.

This study presents several caveats. First, our cohort is not representative of the UK population. For instance, the majority of COVID-19 related deaths in the UK took place in care homes ^49^. However, only six patients included in our study were reported to live in long-term care facilities, and none of them tested positive for COVID-19. Therefore, our analysis of patients affected by chronic neurological diseases only included patients living in domestic residences where rapid changes in social behaviour and domestic care may have affected distinct patient groups differently. This feature might have led us to overestimate the effect of chronic neurological illness on the risk of COVID-19 infection and mortality and further investigation is needed to confirm the generalisability of our results. In addition, it is important to recognise the indirect effects of the current pandemic on AD patients. Because elderly individuals are at increased risk of severe outcomes from COVID-19 infection, government guidelines recommend the isolation of these individuals and limited contact with their family members ^50^. However, social activities as well as time spent with other people is generally considered to help prevent cognitive decline in the elderly ^51^. Therefore, it is plausible that isolation, albeit necessary, may lead to increased stress and cognitive decline among AD patients ^52^. In turn, these behavioural difficulties may exacerbate underlying neurological illness contributing to higher rates of hospitalisation and a higher risk of COVID-19 infection and mortality ^52^. While these effects are challenging to measure in the midst of a pandemic, large-scale retrospective studies will reveal the full range of implications of current isolation measures on AD and COVID-19. Finally, several potentially relevant comorbidities, such as kidney disease, previous myocardial infarction, and stroke, were not included in data collection. Future studies should validate our results by examining the effect of additional comorbidities as well as other factors that we were unable to examine, including access to personal protective equipment, employment, exposure to environmental hazards and the effect of living in care home facilities.

In conclusion, our results support detailed analyses of the biological mechanisms underlying disease-specific vulnerability to SARS-CoV-2 among patients with neurological illness. Improved knowledge of these factors is critical to develop appropriate strategies to protect clinically vulnerable patients affected by neurological disease during this pandemic.

## Data Availability

Code available in GitHub

https://m1gus.github.io/AD_PD_COVID19/

## Abbreviations

AD: Alzheimer’s disease
COPD: Chronic obstructive pulmonary disease
COVID-19: Coronavirus disease-19
PD: Parkinson’s disease
SARS-CoV-2: Severe acute respiratory syndrome coronavirus 2

## AUTHORS’ CONTRIBUTIONS

MT and YY designed the study. YY analysed the data. MT, YY, RP, NSL and LMM wrote the manuscript. NSL and LMM supervised the project. LMM acquired the funding.

## ACKNOWLEDGEMENTS

We are grateful to all the staff members with critical functions in administration, operations and logistics at the MRC Toxicology Unit during the present crisis. We thank the UK Biobank participants and coordinators for the dataset. We also thank Dr Giorgio Fedele for insightful discussions.

